# Projected Trends of Obesity throughout the Life Course According to Sex, Race, and Birth Cohorts in the United States

**DOI:** 10.1101/2024.10.15.24315456

**Authors:** William D. Hazelton, Peiyun Ni, Matthias Harlass, Anne I. Hahn, Ruiyi Tian, Ann G. Zauber, Iris Lansdorp-Vogelaar, Yin Cao

**Affiliations:** Program in Computational Biology, Fred Hutchinson Cancer Research Center, Seattle, WA, USA; Division of Gastroenterology, Department of Medicine, Massachusetts General Hospital and Harvard Medical School, Boston, MA, USA; Department of Public Health, Erasmus Medical Center, Rotterdam, The Netherlands; Department of Epidemiology and Biostatistics, Memorial Sloan Kettering Cancer Center, New York, NY, USA; Division of Public Health Sciences, Department of Surgery, Washington University School of Medicine, St. Louis, MO, USA; Alvin J. Siteman Cancer Center, Washington University School of Medicine, St. Louis, MO, USA; Division of Gastroenterology, Department of Medicine, Washington University School of Medicine, St. Louis, MO, USA

## Abstract

**Background:** Most research on obesity trends and projections has focused on changes across calendar years. However, as the risk of disease increases with cumulative exposure to obesity, it is crucial to characterize the obesity landscape through a life-course perspective and across birth cohorts.

**Objective:** To enhance the accuracy of obesity epidemic projections throughout life course and across birth cohorts in the US.

**Design:** Cross-sectional and cohort study.

**Setting:** United States

**Participants:** Individuals participated in three National Health Examination Surveys (NHES) from 1959 to 1970 and 18 National Health and Nutrition Examination Surveys (NHANES) from 1971 to 2020.

**Measurements:** Body mass index (BMI) distributions by sex, race, and birth cohort.

**Results:** By leveraging over 40 years of cross-sectional and longitudinal data from nationally representative surveys, we developed models to estimate historical and future BMI distributions in the US for both children and adults throughout their life course. We also calculated life-years of exposure to overweight and obesity, according to sex, race, and birth cohort. Our findings reveal significant increases in these metrics among birth cohorts since 1965 and highlight differential trends by sex and race for the 1965, 1985, and 2005 cohorts.

**Limitations:** Assumption that model parameters will hold in the future.

**Conclusion:** Our approach significantly expands upon previous models by projecting life course with continuous BMI distributions informed by longitudinal trajectories, explicitly accounting for variations in birth cohorts.

**Primary Funding Source:** National Institutes of Health.

## INTRODUCTION

Overweight and obesity are major public health problems in the United States. The prevalence of obesity (body mass index (BMI) of 30 kg/m^2^ and greater) has more than tripled from 13% in 1960-1962^1^ to 42% in 2017-2020,^2^ while the prevalence of overweight (BMI between 25 and 29.9 kg/m^2^) remained high, from 32% in 1960-1962 to 30% in 2017-2018.^1^ A greater BMI in mid-adulthood has been causally linked with higher risks of heart diseases,^3^ diabetes,^4^ and 13 types of cancers.^5^ Emerging evidence supports that early-life adiposity may also exert a profound and independent impact on these diseases, independent of adulthood adiposity.^6,7^ Prolonged exposure to adiposity is hypothesized to be a key driver of the rising rates of cardiometabolic disorders^8^ and obesity-related cancers^9^ among younger adults. However, a comprehensive understanding of the trends of the obesity epidemic throughout the life course is lacking.

Obesity trends in the past varied significantly across different sexes, races/ethnicities, and birth cohorts. While non-Hispanic black men had no significant increase in obesity after 2005-2006, with a prevalence of 41% in 2017-2018,^10^ non-Hispanic black women had the highest obesity prevalence among all sex and racial subgroups for three decades, rising from 37% in 1988-1994 to 57% in 2017-2018.^11^ Additionally, more recent birth cohorts are experiencing higher obesity prevalence at younger ages, resulting in longer lifetimes with obesity.^12^ For example, individuals born between 1940 and 1944 reached an obesity prevalence of at least 20% by ages 50 to 54, whereas those born between 1975 and 1979 achieved this level much earlier, by ages 25 to 29.^13^ However, these birth cohort differences have not been incorporated into existing obesity projection models, limiting their ability to fully capture lifetime adiposity exposures across diverse sex and racial populations.

To enhance the accuracy of obesity epidemic projections throughout life course and across birth cohorts, we developed an advanced model that forecasts BMI distributions for both children and adults throughout their lifetimes. This model integrates extensive, longitudinal datasets from the National Health and Nutrition Examination Surveys (NHANES) and National Health Examination Surveys (NHES), spanning 1959 to 2020. Our approach significantly expands upon previous models, which were based on BMI categories,^14–16^ by projecting continuous BMI distributions informed by longitudinal trajectories, explicitly accounting for variations birth cohorts.

## METHODS

### Study population

This study of historical and projected US trends in BMI is based on a series of nationally representative US surveys, including three NHES surveys conducted from 1959 to 1970 and eighteen NHANES surveys conducted from 1971 to 2020. Administered by the National Center for Health Statistics (NCHS), these surveys assess the health of the US population by collecting data on BMI and other factors through structured interviews, biometric measurements, and physical examinations by trained medical technicians. Most of the NHES and NHANES surveys were cross-sectional. However, NHES included a short-term longitudinal follow-up survey of children spanning the years 1963-1970, and NHANES included a sequence of four longitudinal follow-up surveys of adults spanning the years 1971-1992, with NHANES-I as baseline followed by four studies, NHEFS-1 to NHEFS-4 (**Figure 1**).^17^

**Figure 1:**
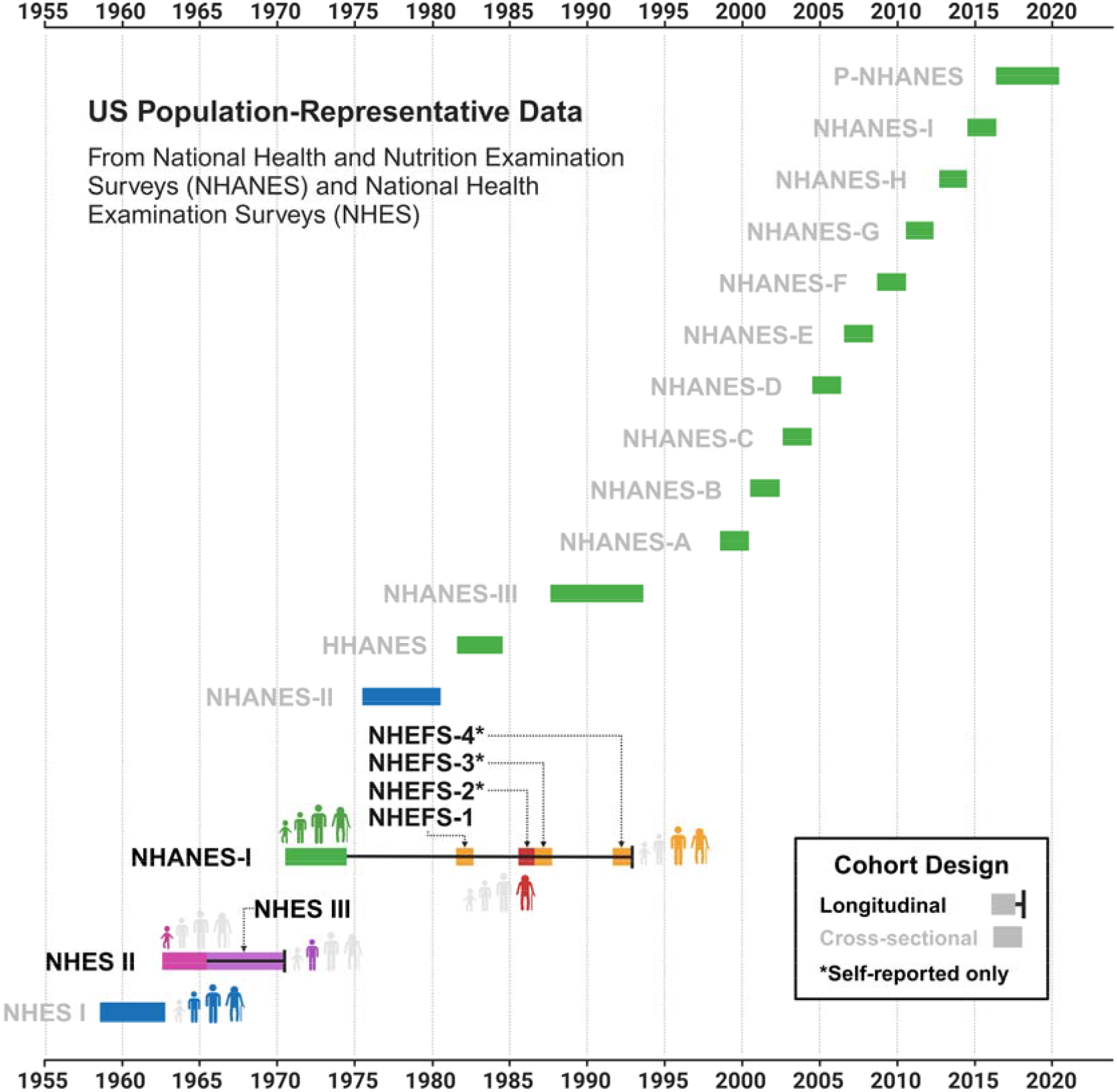
Illustration of national surveys used in the study. NHANES, National Health and Nutrition Examination Survey; P-NHANES, pre-pandemic NHANES; HHANES, Hispanic Health and Nutrition Examination Survey; NHEFS, NHANES I Epidemiologic Follow-up Study; NHES, National Health Examination Survey

The cross-sectional BMI data from NHES and NHANES were used in this study to estimate historical BMI distributions in the US by age, year (or birth cohort), race (non-Hispanic black, non-Hispanic white), and sex, while the longitudinal surveys were used to project BMI trends into the future based on recent trends (under specific assumptions about how trends may continue), and to simulate individual BMI histories for individuals by birth cohorts that collectively match historical BMI trends and future projections. We did not include the Hispanic population due to a lack of longitudinal data.

### Measurement of BMI

Weight and height were retrieved from medical examination records for all NHES and NHANES cross-sectional studies, the longitudinal follow-up in NHES, and the first NHANES follow-up (NHEFS-1) providing 10-year follow-up from baseline (NHANES-I). We also utilized 20-year follow-up (NHEFS-4), but these data were self-reported. BMI was estimated by weight in kilograms (kg) divided by the square of the person’s height in meters. For adults, we used the US Centers for Disease Control (CDC) categorization of overweight (BMI between 25 and 29.9 kg/m^2^) and obesity (BMI ≥ 30 kg/m^2^). For children and adolescents, we used age- and sex-specific 2000 CDC BMI-for-Age growth charts to define underweight as BMI < 5th percentile, healthy weight as BMI between the 5^th^ and 85th percentile, overweight as BMI between the 85th and 95th percentile, and obesity as BMI at or above the 95^th^ percentile. These growth charts were based on data from NHES, NHANES-I, NHANES-II, and NHANES–III.

### Statistical analysis

We estimated US population representative BMI distributions from each NHES and NHANES survey by age, survey year, race, and sex based on individual BMI measurements and individual medical exam sampling weights from each survey. The weighted BMI samples were used to estimate continuous age-specific BMI probability density functions (PDF)s using weighted Gaussian kernel density estimation (KDE) methods. Annual age-specific PDFs between 1960-2020 were then generated using linear interpolation by percentile between successive surveys.

Age-specific longitudinal trends for BMI were estimated based on interpolation across NHANEs longitudinal data by percentile with 10 and 20 years of follow-up. These longitudinal trend estimates were used to extend BMI distributions for surveys that ended before age 85 using matching functions by BMI percentile to extend the last available survey data, and to project recent birth cohorts forward in time while assuming that trends are exponentially damped (with a 20-year time constant) during projection into the future. Individual BMI histories were calculated based on longitudinal transition probabilities between 10 BMI categories using 10- and 20-years of follow-up, while adjusting for cohort trends specific to birth-cohort and age. Final BMI was calculated by sampling from the age- and cohort-specific BMI distribution of the final BMI category.

We used the 1965, 1985 and 2005 birth cohorts to demonstrate the impact of age and cohort on BMI distribution for four groups jointly defined by sex and race (non-Hispanic Whites and non-Hispanic Blacks), forecasting to age 85. In addition, we tested for statistical differences between the age, sex, and race-specific BMI distributions of the 1965 and 1985 birth cohorts using the Anderson-Darling test on samples of 5000 individuals from each distribution. The individual BMI simulator was used to estimate life-course exposures to overweight and obesity among children and adults for these cohorts. The data analysis and visualization were conducted using R version 4.4.0 (R Foundation for Statistical Computing, Vienna, Austria; http://www.R-project.org/).

## RESULTS

### BMI trajectories by birth cohorts

The BMI distributions differed notably by race, sex, and between birth cohorts, particularly for younger age groups (**Table 1; Figures 2,3**). We used the BMI distributions of the birth cohorts from 1965, 1985, and 2005 as examples of the differential age and cohort effects on BMI distribution.

**Table 1:** Median Body Mass Index (BMI, kg/m^2^) of 1965 and 1985 birth cohorts by age, sex, and race. BMI, Body Mass Index; IQR, interquartile range

| Non-Hispanic Black Females |  |  |  |  | Non-Hispanic Black Males |  |  |  |  |
| --- | --- | --- | --- | --- | --- | --- | --- | --- | --- |
| Age | Birth cohort |  | % Change | p-value* | Age | Birth cohort |  | % Change | p-value* |
|  | 1965 | 1985 |  |  |  | 1965 | 1985 |  |  |
|  | Median BMI, (IQR) | Median BMI, (IQR) |  |  |  | Median BMI, (IQR) | Median BMI, (IQR) |  |  |
| 15 | 20.8<br>(18.1-24.1) | 23.2<br>(19.3-28.5) | 11.4 | < .001 | 15 | 20.2<br>(17.7-22.9) | 21.6<br>(18.3-25.9) | 6.9 | < .001 |
| 25 | 25.0<br>(21.0-29.9) | 30.5<br>(23.6-38.3) | 22.1 | < .001 | 25 | 24.7<br>(21.5-28.6) | 26.3<br>(21.2-32.2) | 6.4 | < .001 |
| 35 | 28.7<br>(22.7-35.7) | 31.4<br>(25.0-38.7) | 9.5 | < .001 | 35 | 26.3<br>(21.9-31.4) | 28.0<br>(23.0-34.3) | 6.4 | < .001 |
| 45 | 32.1<br>(25.6-39.6) | 31.6<br>(25.4-37.4) | -1.7 | < .001 | 45 | 29.9<br>(24.3-36.1) | 31.3<br>(26.5-36.0) | 4.4 | < .001 |
| 55 | 33.5<br>(26.7-41.4) | 31.6<br>(25.9-36.9) | -5.6 | < .001 | 55 | 29.1<br>(24.1-34.5) | 29.3<br>(24.7-34.1) | 0.4 | .008 |
| 65 | 31.1<br>(26.2-36.0) | 30.8<br>(26.1-35.6) | -0.7 | .039 | 65 | 28.0<br>(23.0-33.6) | 27.9<br>(22.9-33.3) | -0.4 | .459 |
| 75 | 28.7<br>(23.5-34.3) | 28.5<br>(23.4-34.0) | -0.6 | .098 | 75 | 28.2<br>(23.9-33.1) | 28.1<br>(23.8-32.9) | -0.3 | .008 |
| Non-Hispanic White Females |  |  |  |  | Non-Hispanic White Males |  |  |  |  |
| Age | Birth cohort |  | % Change | p-value* | Age | Birth cohort |  | % Change | p-value* |
|  | 1965 | 1985 |  |  |  | 1965 | 1985 |  |  |
|  | Median BMI, (IQR) | Median BMI, (IQR) |  |  |  | Median BMI, (IQR) | Median BMI, (IQR) |  |  |
| 15 | 20.2<br>(18.1-22.7) | 21.1<br>(18.1-25.0) | 4.5 | < .001 | 15 | 20.2<br>(18.1-22.8) | 20.8<br>(17.8-24.4) | 2.9 | < .001 |
| 25 | 22.4<br>(19.7-26.1) | 24.9<br>(20.7-30.4) | 11.4 | < .001 | 25 | 23.9<br>(21.3-27.0) | 26.0<br>(21.8-30.2) | 8.6 | < .001 |
| 35 | 26.4<br>(21.2-33.0) | 28.6<br>(22.4-36.8) | 8.6 | < .001 | 35 | 26.4<br>(23.0-30.4) | 29.5<br>(24.3-35.4) | 11.7 | < .001 |
| 45 | 25.8<br>(20.8-32.0) | 28.1<br>(23.0-33.9) | 8.8 | < .001 | 45 | 27.8<br>(23.9-32.4) | 28.9<br>(25.2-33.2) | 4.0 | < .001 |
| 55 | 28.6<br>(23.0-35.2) | 28.2<br>(23.0-34.1) | -1.3 | < .001 | 55 | 29.1<br>(24.8-33.9) | 29.2<br>(25.1-33.8) | 0.4 | .001 |
| 65 | 28.9<br>(23.5-35.0) | 28.6<br>(23.3-34.4) | -1.0 | < .001 | 65 | 28.6<br>(24.4-33.4) | 28.6<br>(24.4-33.3) | -0.2 | .962 |
| 75 | 29.5<br>(25.1-34.2) | 29.4<br>(25.1-34.0) | -0.3 | .002 | 75 | 28.4<br>(24.9-32.2) | 28.4<br>(24.9-32.2) | 0.0 | .364 |

\* We used the Anderson-Darling test to compare the complete BMI distributions between birth cohorts. The test was applied to random samples (n = 5000) from the probability density function of each age, sex, race, and birth cohort-specific BMI distribution.

**Figure 2.**
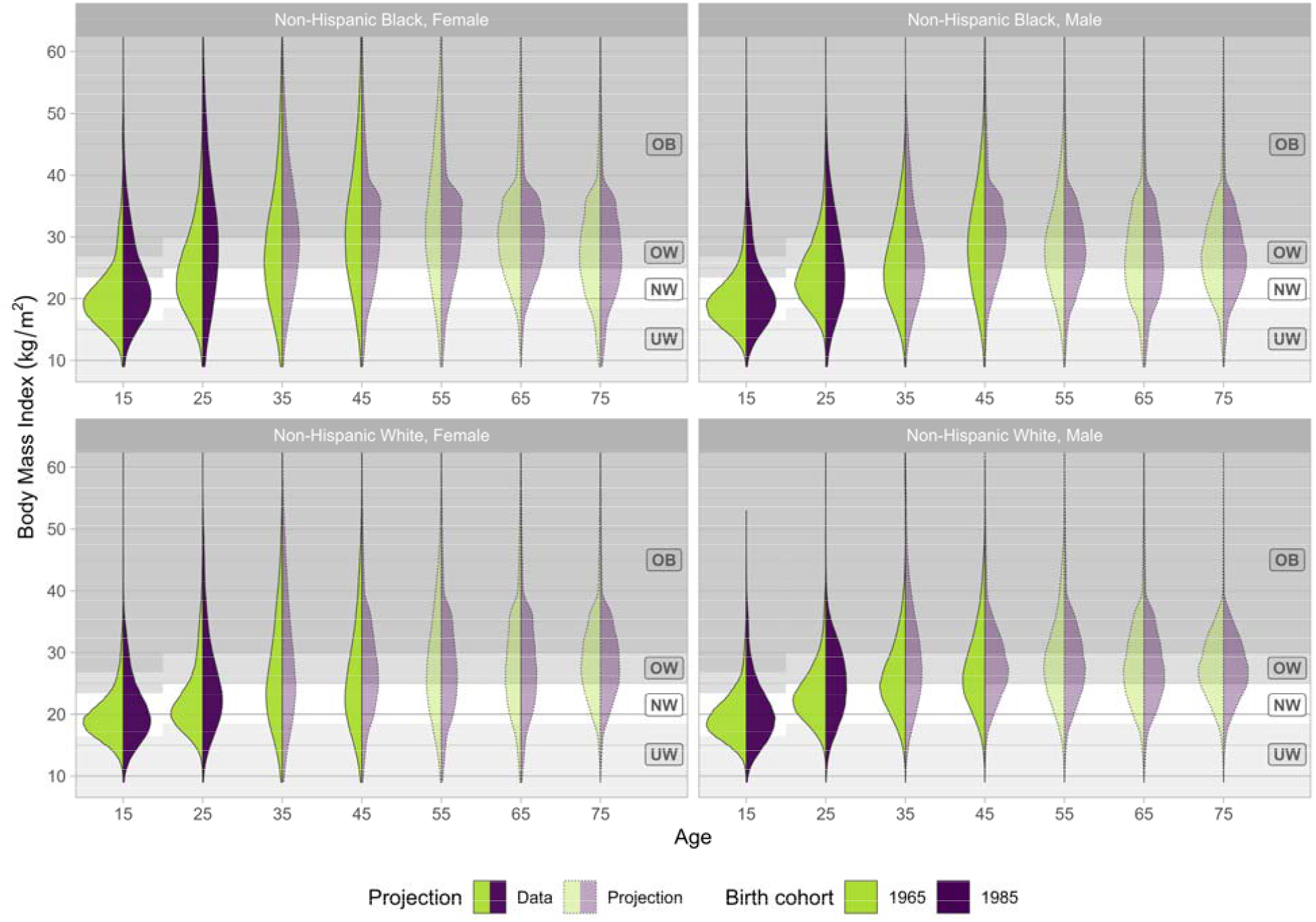
Body Mass Index (BMI, kg/m2) distributions by age, sex, race, and birth cohort. Solid areas represent distributions using observed data and shaded areas represent distributions using future projections. OB, obesity; OW, overweight; NW, normal weight; UW, underweight.

**Figure 3.**
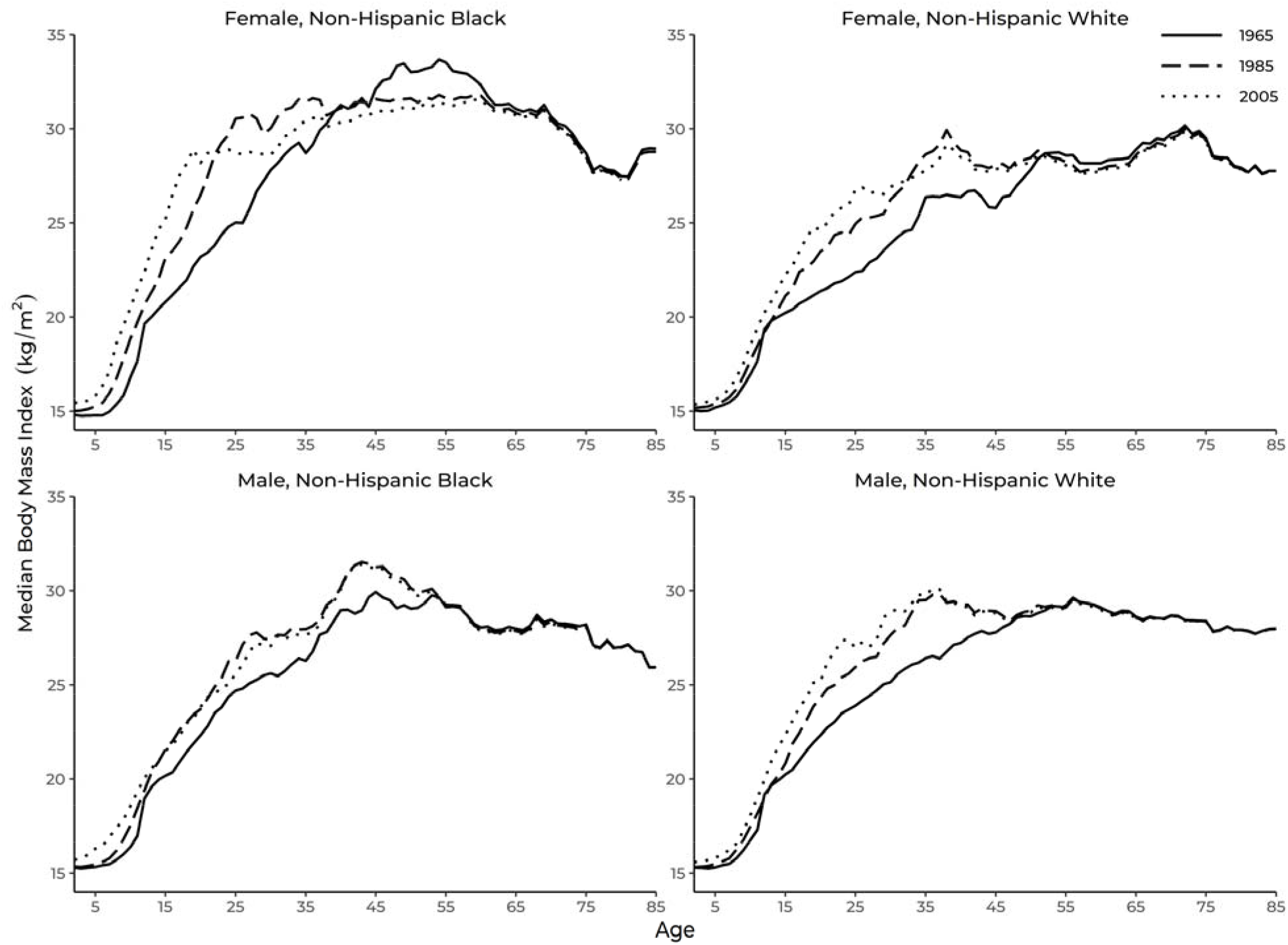
Median Body Mass Index (BMI, kg/m2) distributions by age, sex, race for three birth cohorts.

The median BMI increased for all race and sex groups from birth until ages 35 to 55 and then declined in later years (**Table 1; Figure 3**). Non-Hispanic Black females had the highest median BMI among all subgroups for ages below 35. In 1965, 1985, and 2005, the proportion of obese non-Hispanic Black females at age 35 was 45%, 56%, and 52%, respectively (**Figure 4**). Non-Hispanic white females had the lowest median BMI below age 35 for the 1965 and 1985 birth cohorts, while non-Hispanic Black males had the lowest median BMI below age 35 for the 2005 birth cohort. The proportion of obese non-Hispanic white females at age 35 in 1965 and 1985 was 35% and 45%, and 38% of non-Hispanic Black males were obese in 2005 at age 35 (**Figure 4**).

**Figure 4.**
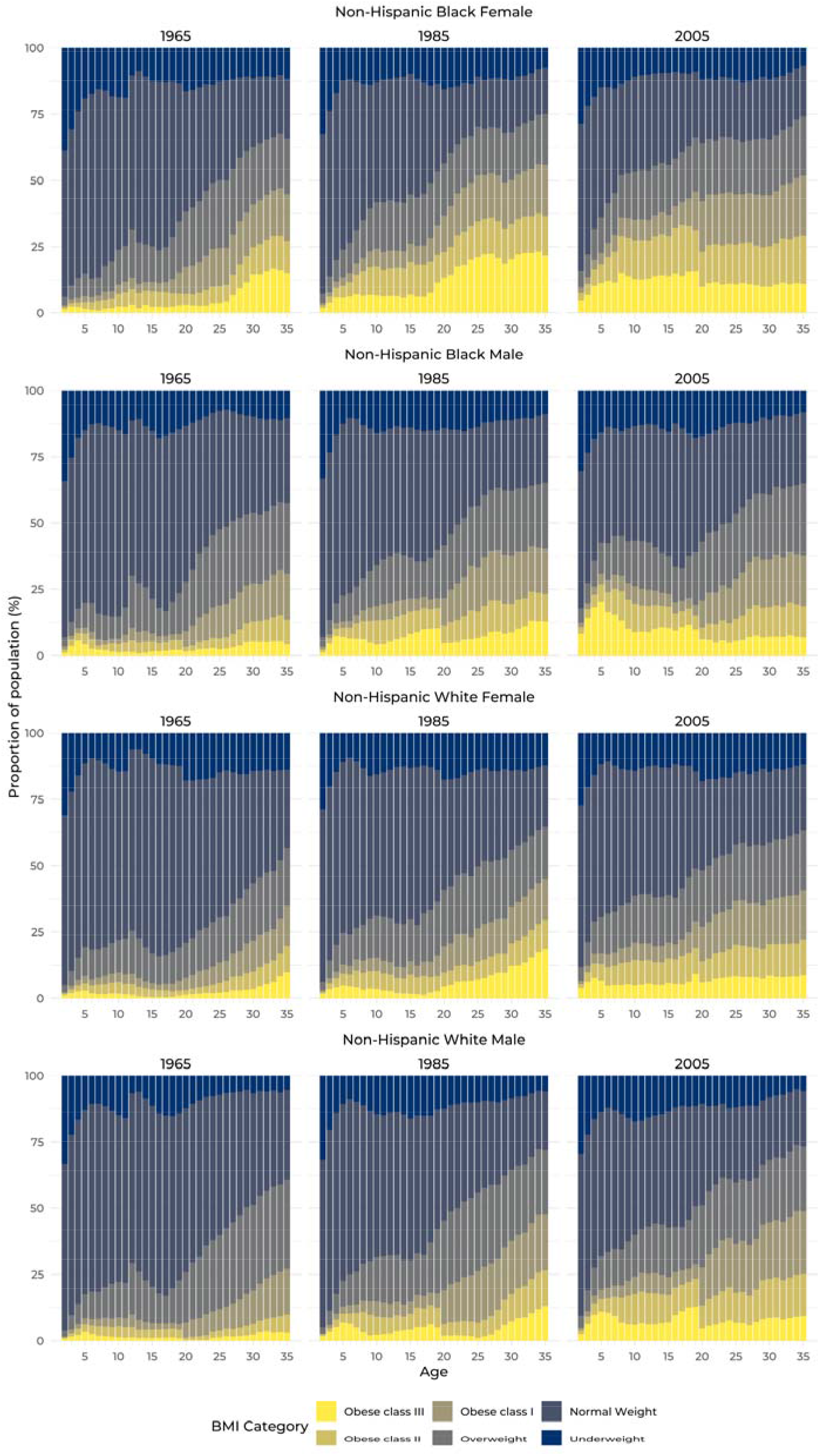
Proportion of Body Mass Index (BMI) categories in young ages by sex, race, and birth cohort.

The comparison between the 1965 and 1985 birth cohorts showed a clear increase in median BMI among younger individuals (aged 15-35), consistent across all sexes and races. Additionally, statistical testing revealed that the BMI distributions differ significantly between birth cohorts until age 45 for all sex and race strata **(Table 1)**. The changes in median BMI were greatest at ages 25 and 35, with the highest BMI increase of 22.1% for non-Hispanic Black females and the smallest increase for non-Hispanic Black males (6.4% BMI increase). Moreover, the proportion of obese individuals below age 35 increased across all strata (**Figure 4**). This indicates a notable shift in the body composition of younger adults over the 20-year period, with differential trends by sex and race.

The trends for the older age groups (45-75 years) were more variable across different sex and race strata (**Table 1**). The median BMI in non-Hispanic Black females over the age of 45 years decreased slightly from the 1965 to 1985 birth cohort but remained similar in non-Hispanic Black males. However, the full BMI density distribution (**Figure 2**) indicated a higher proportion of non-Hispanic Black females with BMIs between 30-35 kg/m2 and a lower proportion with BMIs greater than 40 kg/m2. The median BMI at ages 55 and 65 changed by - 5.6% and -0.7% in non-Hispanic Black females and by 0.4% and -0.4% in non-Hispanic Black males, respectively. A similar pattern was observed for non-Hispanic White females, where the median BMI increased for ages 15 through 45 before slightly decreasing in the older age groups. In contrast, in non-Hispanic White males, the median BMI increased across all age groups, remaining almost constant at ages 65 and 75 between the two birth cohorts.

## DISCUSSION

In this study, we estimated historical and future projections of BMI distributions in the US by age, year, race, and sex for children and adults throughout the life course using cross-sectional and longitudinal data from NHANES and NHES. While previous studies have modeled US population trends for BMI by category, we instead modeled continuous BMI distributions by age and year for different birth cohorts to project future BMI trends by sex and race. We also calculated life-years of exposure to overweight and obesity among children aged 5-19 and separately among adults aged 20-85 by race, sex, and birth cohort. We found significant increases in these metrics among the birth cohorts since 1965 and highlighted the differential trends by sex and race for the 1965, 1985, and 2005 birth cohorts.

Previous BMI projections from 2010 through 2030 were largely based on historical data dating back to 1970-2008.^14,15,18^ While these projections unanimously showed that all sex and racial/ethnic groups were predicted to have rising obesity trends, black women were projected to have the highest obesity prevalence, consistent with current trends.^14,18^ Current projection studies leveraged more recent data, but many did not incorporate the potential impact of birth cohorts^14–16,19^ or include young children and adolescents.^15,16,18^ The obesity trends in young children and adolescents should be explored, as obesity in early childhood can often predict long-term obesity,^20–22^ and is also associated with early onset and higher risks of cardiovascular diseases throughout life.^23,24^ Notably, the rise in adolescent obesity from 1999 to 2018 was found to result primarily from climbing obesity rates in Hispanic and non-Hispanic black youth,^10^ but the evaluation of birth cohort effects on obesity trends for specific sex and racial/ethnic groups in children is still lacking.

Longitudinal BMI data provide information on how BMI distributions change by birth cohort as individuals age and calendar years increase. Moreover, they are important for projecting future population trends as well as for simulating individual BMI trajectories. In general, we found that NHANES and NHES data show trends for BMI increases during childhood and adolescence and decreases late in life, while birth-cohort trends for age-specific BMI distributions have shifted to higher values in recent decades, especially among children and adolescents. While the prediction of future BMI trends is uncertain, longitudinal data can help to more accurately 1) extend available survey data to estimate BMI distributions to older ages, 2) project future trends by birth cohort while estimating upper and lower bound projections to quantify uncertainty, and 3) simulate individual BMI histories for use in modeling of cancer and other diseases.

## CONCLUSION

By projecting continuous BMI distributions over the life course—guided by longitudinal trajectories and explicitly accounting for birth cohort variations—our approach significantly advances previous models. This enhanced precision will enable more accurate identification of high-risk populations, facilitating earlier interventions and more effective disease prevention strategies.

## Data Availability

All data produced in the present study are available upon reasonable request to the authors.

https://wwwn.cdc.gov/nchs/nhanes/Default.aspx

